# Community-based hematological reference intervals among apparently healthy adult Eritrean population in Asmara

**DOI:** 10.1101/2023.04.25.23289092

**Authors:** Ahmed O Noury, Omer A. Musa, Elmuiz Gasmalbari, Barakat M Bakhit, Eyasu H Tesfamariam, Daniel M Abraha, Zekarias B Ghebre

**Affiliations:** Department of physiology, Orotta College of Medicine and Health Sciences, Asmara, Eritrea; Department of Physiology, Faculty of Medicine, the National Ribat Uinversity, Khartoum, Sudan; Department of Biochemistry, Faculty of Medicine, omdurman islamic University. Sudan; Department of Statistics, Biostatistics and Epidemiology Unit, College of Science, Eritrean Institute of Technology, Mai Nefhi, Eritrea; Clinical Laboratory scientist in Hematology department, National Health Laboratory of Eritrea, Asmara, Eritrea; Laboratory Technologist at Eritrean Air Force Hospital, Massawa, Eritrea

## Abstract

**Background:** The complete blood count (CBC) is one of the most beneficial biological tests used in routine medical practice. The reference intervals (RIs) of the hematological parameters are of major importance for clinical orientations and therapeutic decisions and it is necessary to establish RIs that are population specific. The objective of this study was to establish population-specific reference intervals for hematological parameters among healthy adult Eritreans.

**Method:** Using a DXH500 analyzer, CBC values were evaluated in samples taken from 401 healthy Eritreans in Asmara city, ranging in age from 18 to 60. For the blood tests, a sample of venous blood was drawn into a tube containing the anticoagulant EDTA. Data analysis was done using SPSS version 25, and a P value of 0.05 or above was deemed significant. The upper (97.5th percentile) and lower (2.5th percentile) reference interval boundaries with 95% CI were determined using a non-parametric test. The necessity for gender-based reference interval partitioning was determined using the Harris and Boyd Rule.

**Results:** The established 95% reference intervals combined median (2.5th–97.5th percentile) were: that represent both males and females as per the suggestion of Harris and Boyd WBCs, Lymphocytes, Monocytes, Neutrophils, Eosinophils, Basophils, MCV, RDW, RDW-SD and MPV (fl) were 6.3(3.62-11.56×10^3^/μL), 39.53(22.10-60.55 %), 8.67(5.70-13.61 %), 49.32(27.09-69.25 %), 1.19(0.22-7.13%), 0.17(0.02-0.61%), 88.10(79.32-96.07fl), 13.50(12.50-15.90 %), 37.25(33.00-43.29%), and 9.29(7.76-11.51fl) respectively. RBCs, Hb, HCT, MCH, MCHC, and platelets were the parameters that required separate RI. Their respective median (2.5^th^ – 97.5^th^ percentile) for males versus females were 5.40 (4.57-6.21 ×10^6^/μL) versus 4.88 (4.25-5.61×10^6^/μL), 15.66 (13.56-18.13 g/dl) versus 13.50 (11.95-15.68 g/dl, 48 (42.02-53.93%) versus 42.60 (36.40-48.52%), 29.10 (26.02-34.74 pg) versus 28.30 (24.79-31.02 pg), 32.55 (31.60-36.14 g/dl) versus 32.20 (31.10-33.50 14 g/dl) and 273.15 (155.67-399.34) versus 314.35 (113.96-499.55 10^3^/μL).

**Conclusion:** The reference intervals established in this study differ from currently used RIs and thus should be used for the interpretation of laboratory results in diagnosis and safety monitoring in clinical trials in Asmara

## 1. Introduction

Complete blood count (CBC) is the most frequently used laboratory investigation in health care. The most important aspect of laboratory test interpretation is the concept of reference interval (RI), where test values that fall inside the range are considered normal and those occurring outside the range are considered abnormal^(1)^, The concept of population-based RI was introduced in human medicine in 1969 and is defined as the interval within which 95% of a healthy population’s values fall. The most commonly used reference ranges are based on studies conducted in Western countries. ^(2)^ However, these reference values vary depending on a variety of socio-demographic factors. Age, gender, dietary patterns, ethnic differences, and altitude all have an impact on the reference ranges for different groups.^(3)^ The Western RI may not be applicable in most regional settings. To avoid misinterpretation of blood count results, the Clinical and Laboratory Standards Institute (CLSI) recommended that each country establish its own organic standards.^(4)^.

This study was carried out to update the only study of normal blood count values in Asmara in 2018.^(5)^Our research has brought about methodological advancements in sampling and source population selection (household study). The goal of this study is to find out the Hematological reference values in healthy adult Eritreans in Asmara.

## 2. Materials and Methods

### 2.1 Study design

This is a descriptive cross-sectional study

### 2.2 Study site

Asmara is the capital city of Eritrea, a country in the Horn of Africa. It is located in the Central Region of the country. It is the world’s sixth-highest capital by altitude, at an elevation of 2,325 meters (7,628 feet). The city is situated at the tip of an escarpment that separates the Eritrean Highlands from the Great Rift Valley in neighboring Ethiopia. Asmara’s predominant tribe is Tigrinya, and the city has a population of 896,000 people (6)

### 2.3 Volunteer recruitment

The areas from which participants were drawn were chosen using a multistage sampling technique (Stratified sample). The zone was the first-stage sampling unit. Out of several zones in the city, we chose one at random. The second stage involved a random sub-zone selection. We chose one of the zone’s sub-zones at random. The third stage involved a random selection of blocks within the sub-zone. We chose four blocks at random from the sub-zone. The fourth stage involved systematically selecting households from a randomly selected reference point. Following the identification of a household, the zone administrators were contacted, and one employer was assigned to educate the population of the intended blocks about the study’s objectives. Recruitment was stratified into 4 age groups: 18-29, 30-39, 40-49 and 50-60 years.

### 2.4 Reference population

A total of 401 adult Eritreans who belong to various social, ethnic, and professional groups were involved in this cross-sectional study. The participants were targeted for this study according to the Clinical and Laboratory Standards Institute Guidance. Our reference sample consisted of 128 men, with a mean age of 40 years, and 273 women with a mean age of 41 years; the period of study was continued from November 2019 until January 2020.

### 2.5 Ethical clearance

The Eritrean Ministry of Health’s ethics committee granted ethical approval for this study. Before drawing blood, we obtained written informed consent from all participants. The team used a questionnaire to collect anthropometric measurements, demographic data, medical status, medical history, physical activity, and sleeping hours from each blood donor. All participants had their blood pressure, height, and weight measured, and their BMI was calculated.

### 2.6 Blood Collection

An early morning visit was scheduled from 8 a.m. to 12 a.m. A trained phlebotomist collected the blood samples, which were drawn via venipuncture. For a complete blood count, 2 ml of venous blood was collected into tetra-acetic acid (EDTA) vacutainer tubes. Within 5 hours of blood extraction, blood samples were transported to the National Health Laboratory in Asmara, where all analyses were performed at the Hematology Department.

### 2.7 Statistical Analysis

Analysis was done in SPSS (Version 25) after a careful checkup on completeness, cleaning, and editing processes. Extreme values that might greatly affect the result within each gender were identified using the D/R ratio, where D is the absolute difference between an extreme observation (large or small) and the next largest (or smallest) observation, and R is the range (maximum-minimum). The identified extreme values were deleted if D/R≥1/3 (4). Descriptive analysis of the demographic variables, stratified by gender, was performed using mean, median, standard deviation, frequencies, and percent, as appropriate. The Clinical Laboratory Standards Institute /International Federation for Clinical Chemistry (CLSI/IFCC) was employed to compute the reference intervals. As per the standard, a nonparametric method median (IQR), range (Minimum and maximum), combined and separate 95% RIs (2.5th and 97.5th percentiles), 95% CI for the lower limit of RI, and upper limit of RI were computed (using 1000 bootstrapped simple random sampling). Harris and Boyd test vis-à-vis Mann-Whitney U test was performed to determine whether combined or separate RIs are needed (7,8). However, results from Harris and Boyd were finally endorsed. Upon using Harris and Boyd, the statistical Z result was compared with a critical Z* value:Where 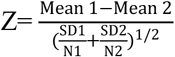 and Z*=3[(N1+N2)/240]1/2. Separate gender specific reference range are needed when Z>Z*.

In order to assess the relationship of the hematological parameters with the socio-demographic and basic background characteristics, spearman rank correlation (for continuous variables), Mann-Whitney u test (categorical variables having dichotomous outcome), and Kruskal-Wallis test (categorical variables having more than two outcomes) was employed. Post-hoc analysis using Bonferroni test was also performed for the results that were found to be significant using Kruskal-Wallis.

Agreement between the current estimated reference intervals and currently used RIs was performed using the Kappa statistic. Interpretation of the Kappa statistic was done using Landis and Koch classification(9).

## 3. Results

### 3.1 Socio-demographic and basic background characteristics

Most of the study participants were females (68.08%). The mean age of the participants was 40.25 (SD=11.68) years. The combined mean (SD) height and weight were 1.62 (SD=0.09) meters and 61.51 (12.11) kilograms, respectively. The mean body mass index was also 23.42 (SD=4.38) kg/m2. The mean (SD) values of SBP and DBP were 118.82 and 76.29 respectively. The mean sleeping hour (per day) was 7.64 hours. The majority (40.1%) of the participants reached secondary education. Married (63.8%) followed by single (25.6%) were predominant among the participants. More than ninety percent of the participants were from the Tigrigna ethnic group (94%) and had three meals per day (98%). 20% of them were found to perform an exercise. A detailed characterization of the study participants stratified by gender is given in Table 1a (continuous variables) and Table 1b (categorical variables).

**Table 1a:**
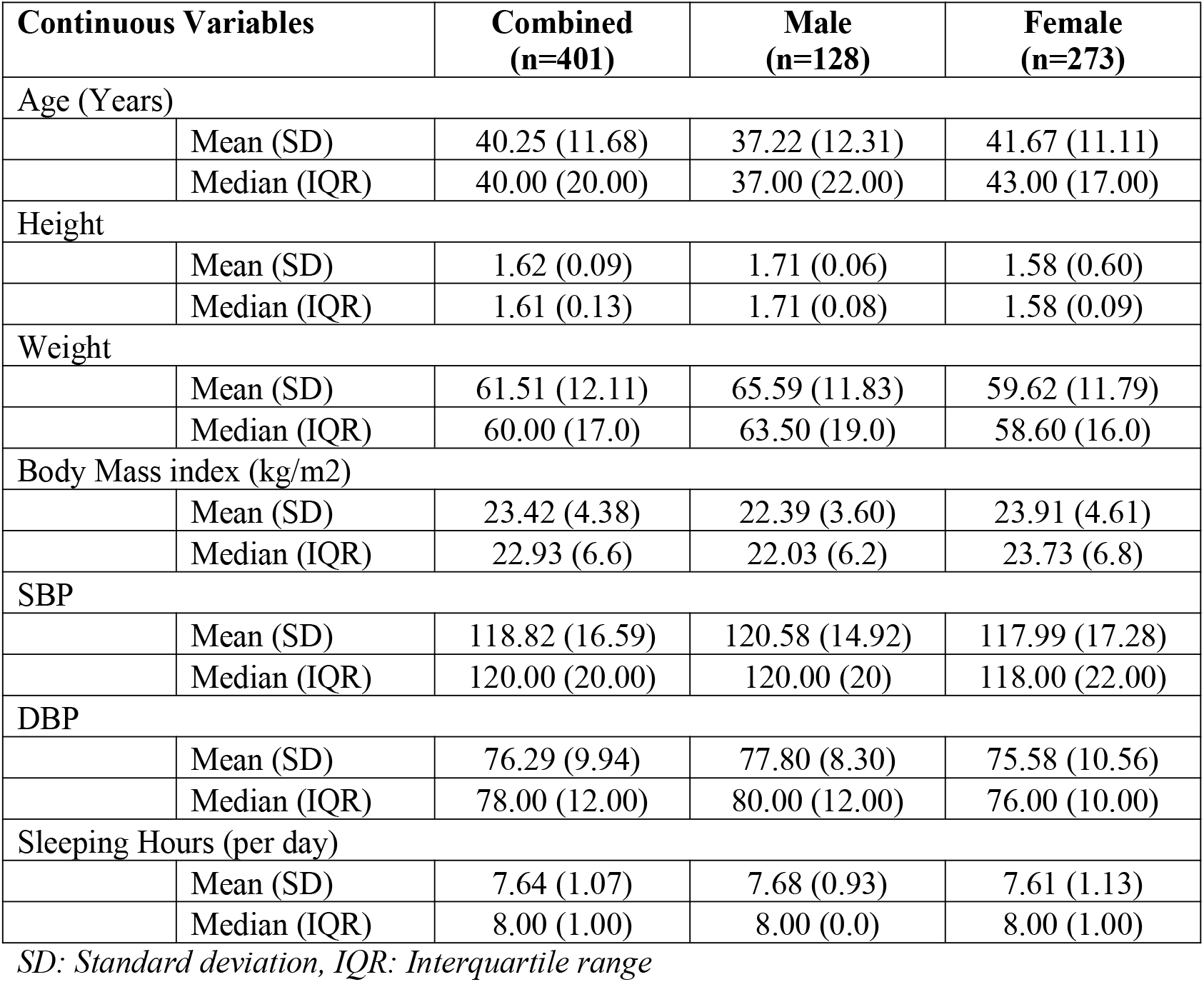
Socio-demographic and basic background characteristics of the study participants for continuous variables stratified by gender (n=401).

**Table 1b:** Socio-demographic and basic background characteristics of the study participants for categorical variables stratified by gender (n=401).

| Categorical Variables |  | Combined<br>(n=401) |  | Male<br>(n=128) |  | Female<br>(n=273) |  |
| --- | --- | --- | --- | --- | --- | --- | --- |
|  |  | n | % | n | % | n | % |
| Highest Educational level |  |  |  |  |  |  |  |
|  | Illiterate | 35 | 8.7 | 0 | 0 | 35 | 12.9 |
|  | Primary | 51 | 12.7 | 6 | 4.7 | 45 | 16.5 |
|  | Junior | 112 | 27.9 | 26 | 20.3 | 86 | 31.6 |
|  | Secondary | 161 | 40.1 | 75 | 58.6 | 86 | 31.6 |
|  | College | 41 | 10.2 | 21 | 16.4 | 20 | 7.4 |
| Marital Status |  |  |  |  |  |  |  |
|  | Married | 256 | 63.8 | 80 | 62.5 | 176 | 64.7 |
|  | Widowed | 17 | 4.2 | 0 | 0 | 17 | 6.3 |
|  | Divorced | 19 | 4.7 | 0 | 0 | 19 | 7 |
|  | Separated | 5 | 1.2 | 0 | 0 | 5 | 1.8 |
|  | Single/Never married | 103 | 25.6 | 48 | 37.5 | 55 | 20.2 |
| Ethnic Group |  |  |  |  |  |  |  |
|  | Tigrigna | 377 | 94 | 119 | 93 | 258 | 94.9 |
|  | Tigre | 11 | 2.7 | 5 | 3.9 | 6 | 2.2 |
|  | Others* | 12 | 2.9 | 3 | 2.4 | 8 | 3 |
| Exercise |  |  |  |  |  |  |  |
|  | Yes | 80 | 20 | 47 | 36.8 | 33 | 12.1 |
|  | No | 321 | 80 | 81 | 63.3 | 240 | 87.9 |
| Number of meals (per day) |  |  |  |  |  |  |  |
|  | Two | 7 | 1.7 | 6 | 4.7 | 1 | 0.4 |
|  | Three | 393 | 98 | 122 | 95.3 | 271 | 99.3 |
|  | Four | 1 | 0.2 | 0 | 0 | 1 | 0.4 |

*Others\* include Afar, Saho, and Blen.*

### 3.2 Parameters that demand partitioned RI and their distribution

The highly recommended test, Harris and Boyd test, was used in determining the need for partitioning of the reference interval by gender (Table 2). The critical value (Z*) was found to be significantly greater than the calculated value (Z) in six out of the 16 assessed hematological parameters, suggesting partitioning of the reference interval by gender is mandatory. The six parameters that need partitioning are RBC, HB, HCT, MCH, MCHC, and PLATELET.

**Table 2:** Hematological parameters and need for partitioning of reference intervals by gender.

| Parameters (Unit) | Harris and Boyd |  | Decision |
| --- | --- | --- | --- |
|  | Z | Z* |  |
| WBCs 103/μL | 0.17 | 3.88 | Combined RI |
| Lymphocytes (%) | 0.46 | 3.88 | Combined RI |
| Monocytes (%) | 3.44 | 3.87 | Combined RI |
| Neutrophils (%) | 1.31 | 3.88 | Combined RI |
| Eosinophils (%) | 1.76 | 3.88 | Combined RI |
| Basophils (%) | 2.21 | 3.88 | Combined RI |
| RBCs $\times 10^6/\mu$L | 12.76 | 3.88 | Separate RI |
| HB (g/dl) | 18.21 | 3.88 | Separate RI |
| HCT (%) | 16.28 | 3.88 | Separate RI |
| MCV (fl) | 3.64 | 3.88 | Combined RI |
| MCH (pg) | 6.33 | 3.87 | Separate RI |
| MCHC (g/dl) | 7.81 | 3.88 | Separate RI |
| RDW (%) | 2.07 | 3.87 | Combined RI |
| RDW-SD | 1.11 | 3.87 | Combined RI |
| Platelets $\times 10^3/\mu$L | 4.89 | 3.87 | Separate RI |
| MPV (fL) | 1.77 | 3.88 | Combined RI |

Z\*, critical value, Z, calculated value, Separate RI are needed only when Z is greater than Z\*; $\dagger S_1^2$ is different from $S_2^2$ .

Though separate reference intervals are needed for the six hematological parameters, as per this study, the reference intervals for all the parameters were computed separately (males and females) as well as the combined data for the sole purpose of making comparisons with other countries’ reference intervals.

Depiction of the patterns and distribution of the parameters that need partitioning (as per Harris and Boyd) by gender are given in Figure 1. It is worth noting that the interval plots are graphed after discarding the outliers found if D/R≥1/3.

**Figure 1:**
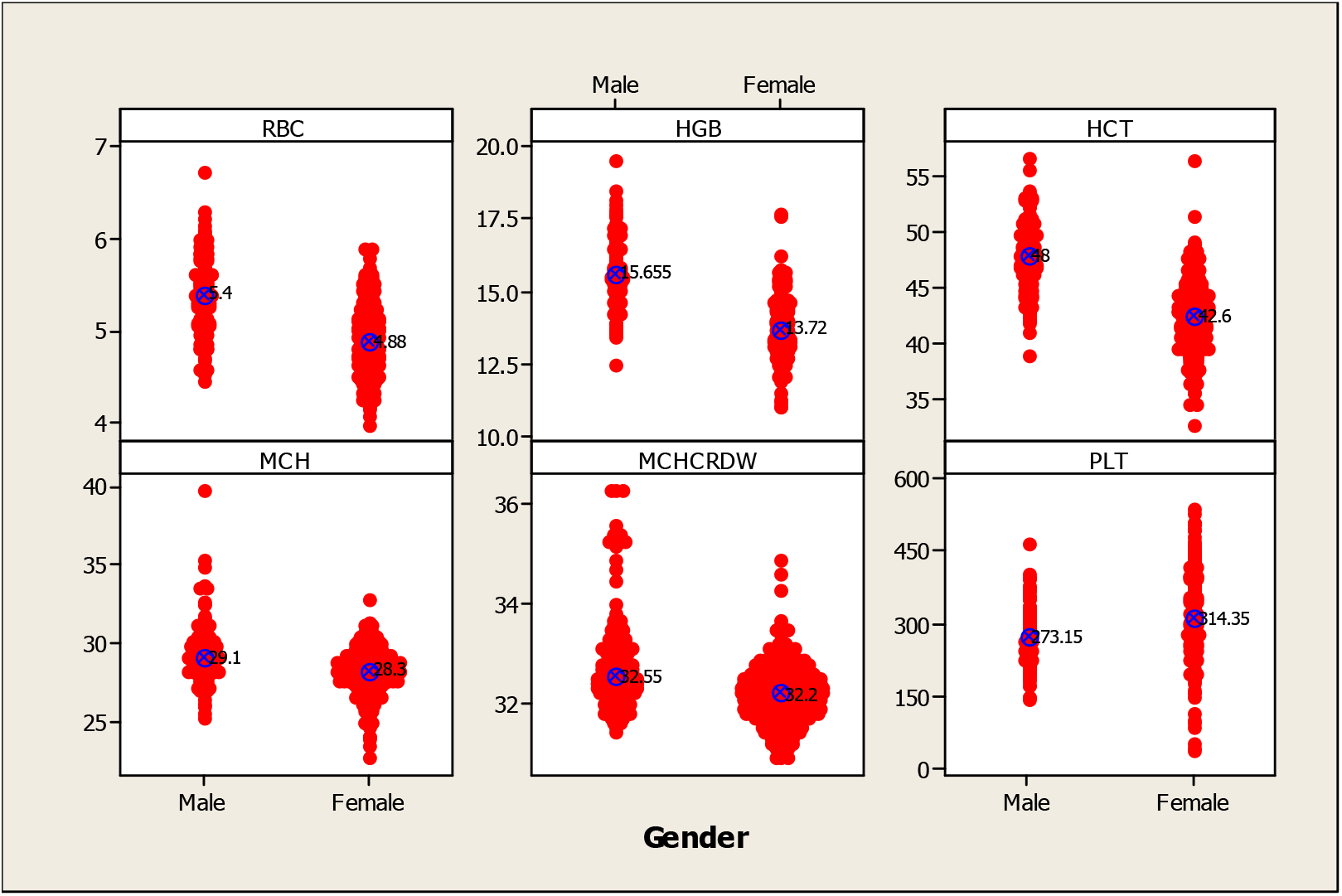
Individual value plots showing location of median and difference of distribution in RBC (×10^12^/L), HB (g/dl), HCT (%), MCH (pg), MCHC (g/dl), and PLATELET(×10^9^/L) among males and females.

### 3.3 Hematological Reference Intervals

Table 3 shows the mean (SD), median (IQR), range (minimum to maximum), 95% reference range (2.5th to 97.5th percentile), 95% CI for the lower limit (2.5th percentile), and 95% CI for the upper limit (97.5th percentile).

**Table 3:**
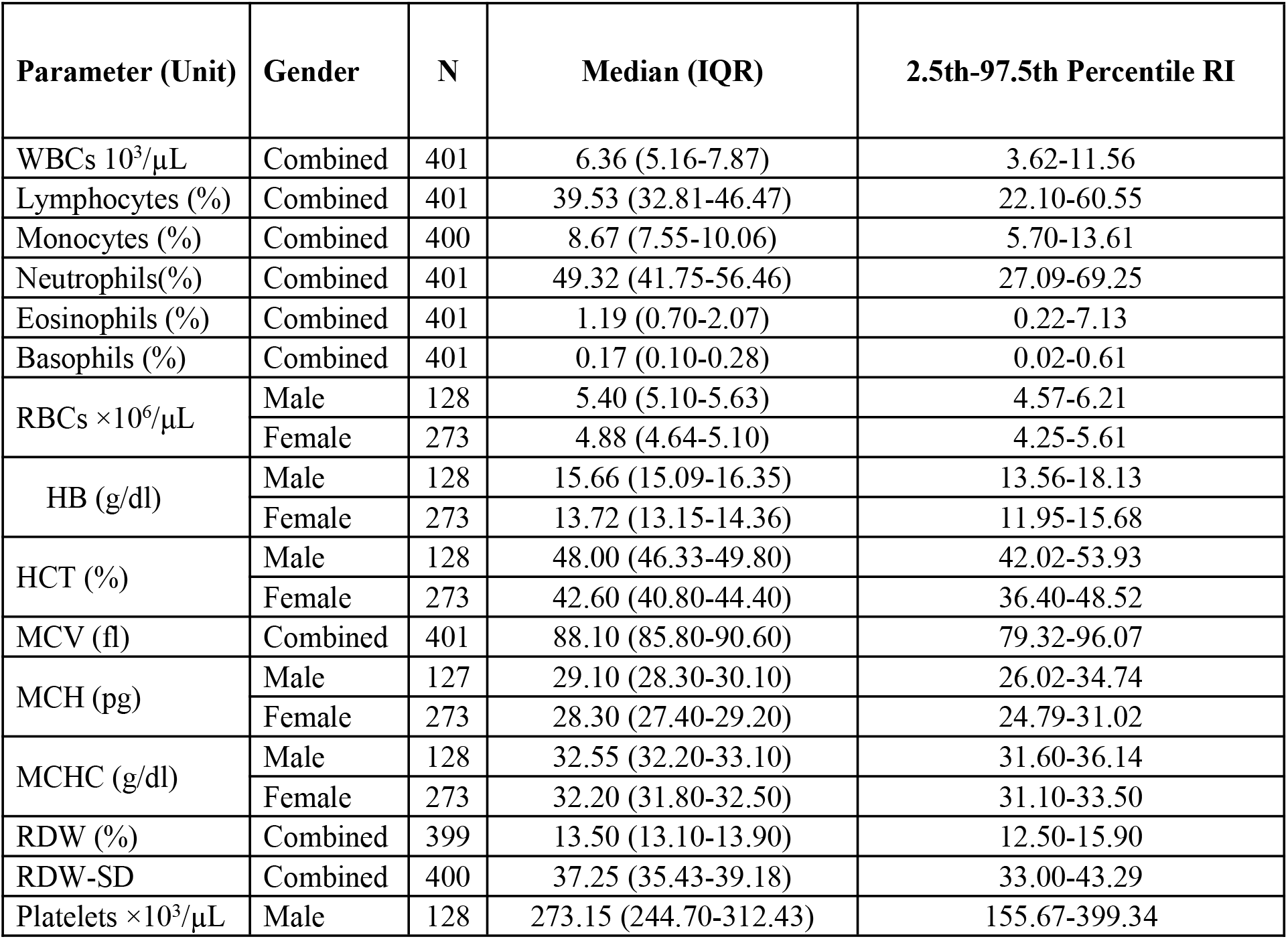
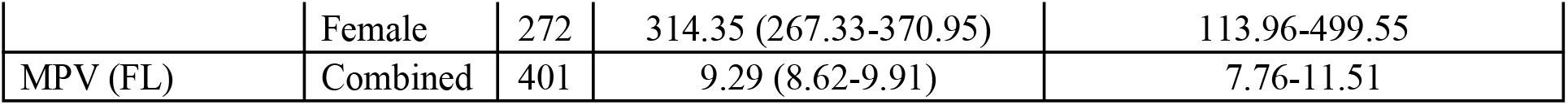
Complete descriptive analysis of the hematological parameters along with the 95% reference range (2.5^th^ to 97.5^th^ percentile) and bootstrapped 95% CI lower limit as well as upper limit.

The combined median (2.5th – 97.5thpercentile) that represents both males and females as per the suggestion of Harris and Boyd for WBC, LYMPHOCYTES, MONOCYTES, NEUTROPHILS, EOSIN, BASOPHILS, MCV, RDW, RDW-SD, and MPV (FL) were6.3(3.62-11.56×109/L), 39.53(22.10-60.55 x109/L), 8.67(5.70-13.61 x109/L), 49.32(27.09-69.25 x109/L), 1.19(0.22-7.13×109/L), 0.17(0.02-0.61×109/L), 88.10(79.32-96.07fL), 13.50(12.50-15.90 %), 37.25(33.00-43.29%), and 9.29(7.76-11.51fL) respectively.

The parameters that demand separate reference ranges as per Harris and Boyd are RBC, HB, HCT, MCH, MCHC, and PLATELET. The median (2.5th – 97.5th percentile) for males versus females are 5.40(4.57-6.21×1012/L) versus 4.88(4.25-5.61×1012/L), 15.66(13.56-18.13g/dl) versus 13.72(11.95-15.68g/dl), 48.00(42.02-53.93 %) versus 42.60(36.40-48.52 %), 29.10(25.2-39.8pg) versus 28.30(22.7-32.8pg), 32.55(31.60-36.14g/dl) versus 32.20(31.10-33.50 g/dl) and 273.15(155.67-399.34×109/L) versus 314.35(113.96-499.55×109/L) for RBC, HB, HCT, MCH, MCHC, and PLATELET respectively.

### 3.4 Relationship of the hematological parameters

Spearman’s correlation of the parameters with age revealed that BASOPHILS (r=0.123, p=0.015), RBC (r=-0.156, p=0.002), HB (r=-0.126, p=0.013), and HCT (r=-0.125, p=0.014), RDW (r=0.109, p=0.032), RDW-SD (r=0.160, p=0.002), and PLATELET (r=-0.149, p=0.003) were significantly related (Table 4). With an increase in age, a significant increase in BASOPHILS, RDW, and RDW-SD was observed. However, with increased age, a significant decrease in RBC, HB, HCT, and PLATELET was observed.

**Table 4:**
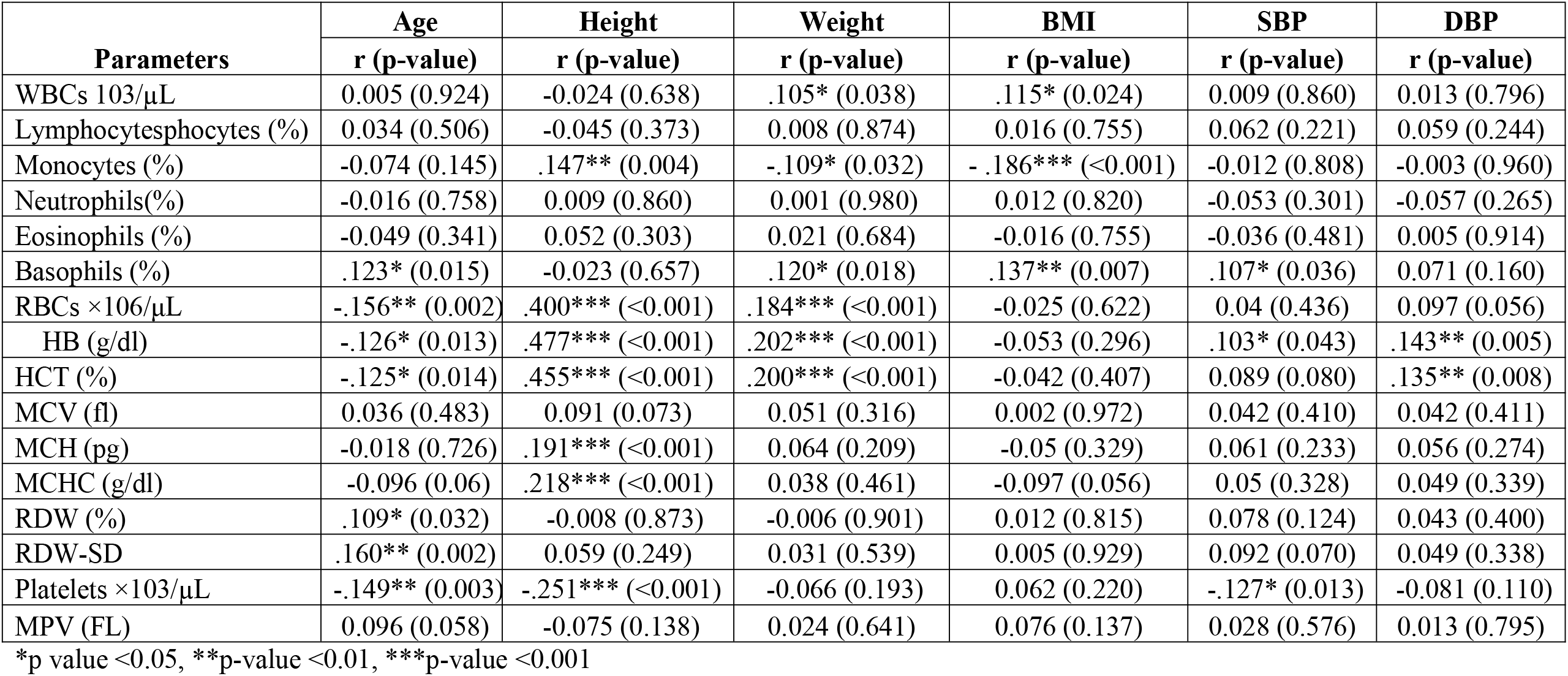
Spearman’s correlation of the hematological parameters with the continuous background variables (n=401)

| Parameters | Age | Height | Weight | BMI | SBP | DBP |
| --- | --- | --- | --- | --- | --- | --- |
|  | r (p-value) | r (p-value) | r (p-value) | r (p-value) | r (p-value) | r (p-value) |
| WBCs 103/μL | 0.005 (0.924) | -0.024 (0.638) | .105* (0.038) | .115* (0.024) | 0.009 (0.860) | 0.013 (0.796) |
| Lymphocytes (%) | 0.034 (0.506) | -0.045 (0.373) | 0.008 (0.874) | 0.016 (0.755) | 0.062 (0.221) | 0.059 (0.244) |
| Monocytes (%) | -0.074 (0.145) | .147** (0.004) | -.109* (0.032) | - .186*** (<0.001) | -0.012 (0.808) | -0.003 (0.960) |
| Neutrophils (%) | -0.016 (0.758) | 0.009 (0.860) | 0.001 (0.980) | 0.012 (0.820) | -0.053 (0.301) | -0.057 (0.265) |
| Eosinophils (%) | -0.049 (0.341) | 0.052 (0.303) | 0.021 (0.684) | -0.016 (0.755) | -0.036 (0.481) | 0.005 (0.914) |
| Basophils (%) | .123* (0.015) | -0.023 (0.657) | .120* (0.018) | .137** (0.007) | .107* (0.036) | 0.071 (0.160) |
| RBCs $\times 106/\mu$L | -.156** (0.002) | .400*** (<0.001) | .184*** (<0.001) | -0.025 (0.622) | 0.04 (0.436) | 0.097 (0.056) |
| HB (g/dl) | -.126* (0.013) | .477*** (<0.001) | .202*** (<0.001) | -0.053 (0.296) | .103* (0.043) | .143** (0.005) |
| HCT (%) | -.125* (0.014) | .455*** (<0.001) | .200*** (<0.001) | -0.042 (0.407) | 0.089 (0.080) | .135** (0.008) |
| MCV (fl) | 0.036 (0.483) | 0.091 (0.073) | 0.051 (0.316) | 0.002 (0.972) | 0.042 (0.410) | 0.042 (0.411) |
| MCH (pg) | -0.018 (0.726) | .191*** (<0.001) | 0.064 (0.209) | -0.05 (0.329) | 0.061 (0.233) | 0.056 (0.274) |
| MCHC (g/dl) | -0.096 (0.06) | .218*** (<0.001) | 0.038 (0.461) | -0.097 (0.056) | 0.05 (0.328) | 0.049 (0.339) |
| RDW (%) | .109* (0.032) | -0.008 (0.873) | -0.006 (0.901) | 0.012 (0.815) | 0.078 (0.124) | 0.043 (0.400) |
| RDW-SD | .160** (0.002) | 0.059 (0.249) | 0.031 (0.539) | 0.005 (0.929) | 0.092 (0.070) | 0.049 (0.338) |
| Platelets $\times 103/\mu$L | -.149** (0.003) | -.251*** (<0.001) | -0.066 (0.193) | 0.062 (0.220) | -.127* (0.013) | -0.081 (0.110) |
| MPV (fL) | 0.096 (0.058) | -0.075 (0.138) | 0.024 (0.641) | 0.076 (0.137) | 0.028 (0.576) | 0.013 (0.795) |

\*p value <0.05, \*\*p-value <0.01, \*\*\*p-value <0.001

Seven out of sixteen hematological parameters were found to have a significant correlation with height. Positive significant correlation was observed between height and MONOCYTES (r=0.147, p=0.004), RBC (r=0.400, p<0.001), HB (r=0.477, p<0.001), HCT (r=0.455, p<0.001), MCH (r=0.191, p<0.001), MCHC (r=0.218, p<0.001). On the other hand, a decrease in RDW-SD (r=-0.251, p<0.001) was observed with an increase in height.

Weight was found to have a significant correlation with six out of sixteen hematological parameters, viz., WBC, MONOCYTES, BASOPHILS, RBC, HB, and HCT. Out of the six significantly related ones, a negative correlation was observed between weight and MONOCYTES (r=-0.109, p=0.032) only, depicting a decrease in MONOCYTES with an increase in weight. The remaining parameters WBC (r=0.105, p=0.038), BASOPHILS (r=0.120, p=0.018), RBC (r=0.184, p<0.001), HB (r=0.202, p<0.001), and HCT (r=0.200, p<0.001) were positively correlated with weight.

A positive significant increase in BMI (r=0.115, p=0.024) and BASOPHILS (r=0.137, p=0.007) was observed with an increase in BMI. However, a negative correlation was observed between MONOCYTES (r=-0.186, p<0.001) and BMI. The remaining thirteen hematological parameters were not significantly correlated with BMI.

Only three hematological parameters were found to have a significant correlation with SBP, namely, BASOPHILS, HB, and PLATELET. Out of the three parameters, a significant decrease in PLATELET (r=0.127, p=0.013) was observed with an increase in SBP. However, a significant increase in BASOPHILS (r=0.107, p=0.036) and HB (r=0.103, p=0.043) was observed with an increase in SBP.

HB and HCT were significantly related to DBP. A significant positive relationship was observed between both HB (r=0.143, p=0.005) and HCT (r=0.1335, p=0.008) and DBP.

Mann-Whitney U test revealed significantly higher MONOCYTES (p=0.001), EOSIN (p=0.019), RBC (p<0.001), HB (p<0.001), HCT (p<0.001), MCV (p=0.001), MCH (p<0.001), MCHC (p<0.001) among males as compared to females. On the other hand PLATELET was higher among females as compared to males (p<0.001).

### 3.5 Comparison of the hematological parameters with currently used RI

The hematological RIs established in this study were also compared to the currently used hematological RI in Asmara. This was performed by computing the percent agreement, and kappa concordance and results were interpreted as per the Landis and Koch interpretation.

An almost perfect agreement was obtained between the estimated RI and currently used RI in Asmara for HCT (Kappa=0.84, 95% CI: 0.72, 0.97) and PLATELET (Kappa=0.82, 95% CI: 0.70-0.95). Substantial agreement was observed for RBC (Kappa=0.71, 95% CI: 0.51-0.90) and MCV (Kappa=0.70, 95% CI: 0.55-0.84). On the other hand, a fair agreement was observed for HB (Kappa=0.40, 95% CI: 0.27-0.53), MCH (Kappa=0.38, 95% CI: 0.25-0.50), RDW (Kappa=0.440, 95% CI: 0.22-0.58), and WBC (Kappa=0.34, 95% CI: 0.22-0.46). Slight agreement (kappa=0.12, 95% CI: 0.04-0.20) was observed for MCHC.

## Discussion

The purpose of this study was to establish hematological reference ranges in healthy adult Eritreans in Asmara to serve as standards for the interpretation of laboratory results in clinical practice for diagnosis and follow-up, as well as screening in routine healthcare in Asmara. The researchers also wanted to see how age, gender, height, BMI, and systolic and diastolic blood pressure affected hematological variables in healthy adults. A comparison was made between our findings and currently used RIs, A comparison was made between this study and other studies published in literature; the comparison of our finding and other studies of hematological values in different countries were presented in the Tables (6,7,8 and 9). The careful recruitment of healthy individuals with strict exclusion criteria is the study’s main strength. This improves the results’ validity.

**Table 5:** Comparison of the hematological parameters among males and females.

| Parameters | Male | Female | P-value |
| --- | --- | --- | --- |
|  | Md (IQR) | Md (IQR) |  |
| WBCs $10^3/\mu\text{L}$ | 6.29 (3.08) | 6.39 (2.59) | 0.624 |
| Lymphocytesphocytes (%) | 38.66 (12.81) | 40.49 (14.36) | 0.883 |
| Monocytes (%) | 9.11 (2.78) | 8.52 (2.74) | **0.001** |
| Neutrophils(%) | 50.11 (13.28) | 49.00 (16.19) | 0.397 |
| Eosinophils (%) | 1.29 (1.59) | 1.06 (1.25) | **0.019** |
| Basophils (%) | 0.16 (0.18) | 0.18 (0.19) | 0.063 |
| RBCs $\times 10^6/\mu\text{L}$ | 5.40 (0.54) | 4.88 (0.46) | **<0.001** |
| HB (g/dl) | 15.65 (1.26) | 13.72 (1.21) | **<0.001** |
| HCT (%) | 48.00 (3.5) | 42.60 (3.6) | **<0.001** |
| MCV (fl) | 89.00 (4.4) | 87.70 (4.90) | **0.001** |
| MCH (pg) | 29.10 (1.8) | 28.30 (1.8) | **<0.001** |
| MCHC (g/dl) | 32.55 (0.9) | 32.20 (0.7) | **<0.001** |
| RDW (%) | 13.40 (0.9) | 13.50 (0.7) | 0.135 |
| RDW-SD | 37.20 (3.8) | 37.20 (3.7) | 0.481 |
| Platelets $\times 10^3/\mu\text{L}$ | 273.15 (67.7) | 314.50 (103.6) | **<0.001** |
| MPV ( fL) | 9.24 (1.18) | 9.31 (1.36) | 0.162 |

\*p value <0.05, \*\*p-value <0.01, \*\*\*p-value <0.001

**Table 6:** Agreement between the estimated and currently used RIs for selected parameters.

|  |  | Normal classification using estimated RI |  | Percent agreement | Kappa (95% CI) | Landis and Koch interpretation |
| --- | --- | --- | --- | --- | --- | --- |
| Parameters | Normal classification using MoH | No n(%) | Yes n(%) |  |  |  |
| RBC*10<sup>12</sup>/L | No | 10 (2.5) | 0 (0) | 98.00% | 0.71 (0.51-0.90) | Substantial |
|  | Yes | 8 (2.0) | 383 (95.5) |  |  |  |
| HB (g/dl) | No | 19 (4.7) | 48 (12.0) | 88.00% | 0.40 (0.27-0.53) | Fair |
|  | Yes | 0 (0) | 334 (83.3) |  |  |  |
| HCT (%) | No | 17 (4.2) | 6 (1.5) | 98.50% | 0.84 (0.72-0.97) | Almost perfect |
|  | Yes | 0 (0) | 384 (94.3) |  |  |  |
| MCV (fl) | No | 20 (5.0) | 16 (4.0) | 96.00% | 0.70 (0.55-0.84) | Substantial |
|  | Yes | 0 (0) | 365 (91.0) |  |  |  |
| MCH (pg) | No | 20 (5.0) | 54 (13.5) | 86.50% | 0.38 (0.25-0.50) | Fair |
|  | Yes | 0 (0) | 327 (81.5) |  |  |  |
| MCHC (g/dl) | No | 11 (2.7) | 100 (24.9) | 74.30% | 0.12 (0.04-0.2) | Slight |
|  | Yes | 3 (0.7) | 287 (71.6) |  |  |  |
| RDW (%) | No | 11 (2.7) | 23 (5.7) | 92.70% | 0.40 (0.22-0.58) | Fair |
|  | Yes | 6 (1.5) | 361 (90.0) |  |  |  |
| WBC*10<sup>9</sup>/L | No | 18 (4.5) | 58 (14.5) | 85.50% | 0.34 (0.22-0.46) | Fair |
|  | Yes | 0 (0) | 325 (81.0) |  |  |  |
| Platelets ×103/μL | No | 20 (5.0) | 7 (1.7) | 98.00% | 0.82 (0.70-0.95) | Almost perfect |
|  | Yes | 1 (0.2) | 373 (93.0) |  |  |  |

*MoH: Ministry of Health*

**Table 7:** Comparison of the hematological parameters referanace interval with international reference interval(10)

| Total reference value<br>Median and 95% reference range (2.5th–95th percentiles) |  |  |  |  |
| --- | --- | --- | --- | --- |
| Parameter | Males |  | females |  |
|  | Present study | International | Present study | International |
| **RBC\*1012/L** | 4.57-6.21(5.39) | 3.80–5.50 (4.7) | 4.25-5.61 (4.89) | 3.60–4.80 (4.2) |
| **HB (g/dl)** | 13.56-18.13 (15.74) | 13.0-17.0 (15) | 11.95-15.68 (13.75) | 12.0-15.0 (13.5) |
| **HCT (%)** | 42.02-53.93 (47.94) | 45(40-50) () | 36.40-48.52 (42.62) | 40(35-45) () |
| **MCV (fl)** | 81.35-96.43 (89.10) | 92(83-101) () | 78.16-95.30 (87.5) | 92(83-101) () |
| **MCH (pg)** | 26.02-34.74 (29.34) | 29(26-32) () | 24.79-31.02 (28.22) | 29(26-32) () |
| **MCHC(%)** | 31.60-36.14 (32.85) | 33(31-35) | 31.10-33.50 (32.21) | 33(31-35) |
| **RDW (%)** | 12.52-14.88 (13.49) |  | 12.50-16.80(13.69) |  |
| **WBC\*109/L** | 3.19-12.29 (6.65) | 4-11 (7.5) | 3.80-11.37 (6.69) | 4-11 (7.5) |
| **Platelets**<br>**×103/μL** | 155.67-399.34 (277.3) | 120–410 (275) | 113.96-499.55 (317.7) | 120–410 (275) |

**Table 8:** Comparison of the hematological parameters reference interval in Asmara with African values.

| Total reference value<br>Median and 95% reference range (2.5th–95th percentiles) |  |  |  |  |  |  |
| --- | --- | --- | --- | --- | --- | --- |
| Parameter | Present study | Sudan<sup>((10,11))</sup> | Ethiopia<sup>(13)</sup> | Ghana<sup>(15)</sup> | Togo<sup>(16)</sup> | Zimbabwean<sup>(17)</sup> |
| **RBC\*10<sup>12</sup>/L** |  |  |  |  |  |  |
| **Males** | 4.57-6.21 (5.39) | 5.08(3.94-6.13) | 5.1 (4.8–5.5) | 3.79–5.96 (4.8) | 3.3–6.4 (5) | 4.4–6.7 (5.5) |
| **females** | 4.25-5.61 (4.89) | 4.52(3.67-5.44) | 4.5 (4.2–4.8) | 3.09–5.30 (4.3) | (3.1–6)4.5 | 3.9–5.9 (4.7) |
| **HB (g/dl)** |  |  |  |  |  |  |
| **Males** | 13.56-18.13 (15.74) | 15.1(11.3-17.4) | 15.3 (14.2–16.5) | 11.3–16.4 (13.9) | 10–18.4 (15.1) | 13.2–18.3 (15.9) |
| **females** | 11.95-15.68 (13.75) | 12.6(9.3-15.6) | 13.1 (12.1–14.3) | 8.8–14.4 (12.3) | 10.3–17.1 (13) | 10.2–15.9(13.5) |
| **HCT (%)** |  |  |  |  |  |  |
| **Males** | 42.02-53.93 (47.94) | 44.5(34.3-56.8) | 46.2 (43.6–49.2) | 33.2–50.5 (42) | 28–54 (43) | 42.0–55.1 (48.5) |
| **females** | 36.40-48.52 (42.62) | 38.1(30.2-48.5) | 39.9 (37.6–43.5) | 26.4–45 (37) | 28–47 (38) | 33.9–48.7 (41.7) |
| **MCV (fl)** |  |  |  |  |  |  |
| **Males** | 81.35-96.43<br>(89.10) | 88.1(73.9-108) | 90.1 (87–93) | 70–98 (87) | 80–99 (85) | 72.8–102 (88) |
| **females** | 78.16-95.30<br>(87.57) | 85.5(70.8-101) |  | 73–96 (86) | 80–95 (84) | 68.8–100 (88) |
| **MCH (pg)** |  |  |  |  |  |  |
| **Males** | 26.02-34.74<br>(29.34) | 29.7(21.6-33.9) | 30 (28.1–31.7) | 22.3–33.6<br>(28.6) | 26–36<br>(29.5) | 22.9–33.5<br>(29.4) |
| **females** | 24.79-31.02<br>(28.22) | 28.2(20.3-32.9) |  | 22.6–33.5<br>(28.6) | 25–37<br>(29.3) | 20.7–32.1<br>(28.4) |
| **MCHC(g/d)** |  |  |  |  |  |  |
| **Males** | 31.60-36.14<br>(32.85) | 33.2(28.0-37.1) | 33.9 (30.5–35.1) | 30.6–36<br>(33.1) | 29–39<br>(35.1) | 29.8–35.4<br>(32.7) |
| **females** | 31.10-33.50<br>(32.21) | 32.6(27.5-36.8) | 33.9 (30.5–35.1) | 30.4–36.5<br>(33.1) | 30–41<br>(35.1) | 29.2–34.3<br>(32.1) |
| **WBC\*10<sup>9</sup>/L** |  |  |  |  |  |  |
| **Males** | 3.19-12.29<br>(6.65) | 4.969 | 6.1 (4.8–7.7) | 3.5–9.2<br>(5.5) | 1.9–10.1<br>(4.1) | 2.8–8.1 (4.6) |
| **females** | 3.80-11.37<br>(6.69) | 5.138 | 6.1 (4.8–7.7) | 3.4–9.3<br>(5.3) | 1.9–10.1<br>(4.1) | 3.3–8.3 (5.2) |
| **Platelets ×103/μL** |  |  |  |  |  |  |
| **Males** | 155.67-399.34<br>(277.34) | 200- 291<br>(245) | 239 (186–293) | 88–352<br>(208) | 120–443<br>(236) | 125–357<br>(229) |
| **females** | 113.96-499.55<br>(317.72) | 243- 347<br>(295) | 239 (186–293) | 89–403<br>(224) | 150–436<br>(247) | 163–431<br>(268) |

**Table 9:**
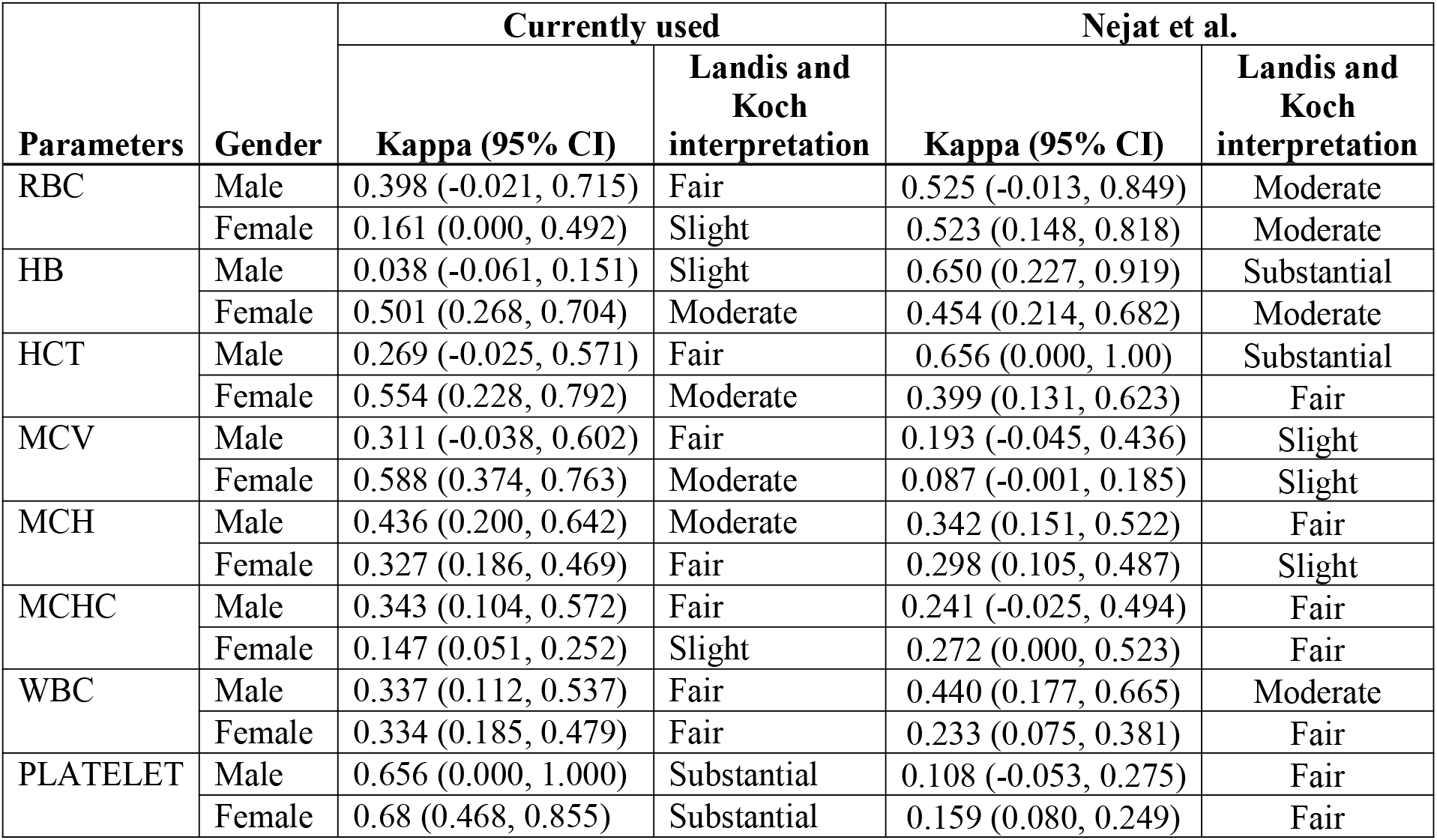
Comparison with the only study done in Asmara.

| Parameters | Gender | Currently used |  | Nejat et al. |  |
| --- | --- | --- | --- | --- | --- |
|  |  | Kappa (95% CI) | Landis and Koch interpretation | Kappa (95% CI) | Landis and Koch interpretation |
| RBC | Male | 0.398 (-0.021, 0.715) | Fair | 0.525 (-0.013, 0.849) | Moderate |
|  | Female | 0.161 (0.000, 0.492) | Slight | 0.523 (0.148, 0.818) | Moderate |
| HB | Male | 0.038 (-0.061, 0.151) | Slight | 0.650 (0.227, 0.919) | Substantial |
|  | Female | 0.501 (0.268, 0.704) | Moderate | 0.454 (0.214, 0.682) | Moderate |
| HCT | Male | 0.269 (-0.025, 0.571) | Fair | 0.656 (0.000, 1.00) | Substantial |
|  | Female | 0.554 (0.228, 0.792) | Moderate | 0.399 (0.131, 0.623) | Fair |
| MCV | Male | 0.311 (-0.038, 0.602) | Fair | 0.193 (-0.045, 0.436) | Slight |
|  | Female | 0.588 (0.374, 0.763) | Moderate | 0.087 (-0.001, 0.185) | Slight |
| MCH | Male | 0.436 (0.200, 0.642) | Moderate | 0.342 (0.151, 0.522) | Fair |
|  | Female | 0.327 (0.186, 0.469) | Fair | 0.298 (0.105, 0.487) | Slight |
| MCHC | Male | 0.343 (0.104, 0.572) | Fair | 0.241 (-0.025, 0.494) | Fair |
|  | Female | 0.147 (0.051, 0.252) | Slight | 0.272 (0.000, 0.523) | Fair |
| WBC | Male | 0.337 (0.112, 0.537) | Fair | 0.440 (0.177, 0.665) | Moderate |
|  | Female | 0.334 (0.185, 0.479) | Fair | 0.233 (0.075, 0.381) | Fair |
| PLATELET | Male | 0.656 (0.000, 1.000) | Substantial | 0.108 (-0.053, 0.275) | Fair |
|  | Female | 0.68 (0.468, 0.855) | Substantial | 0.159 (0.080, 0.249) | Fair |

The median values for MCH, MCHC, and male platelets count are comparable to those of Western populations (Table 7), which is consistent with other African studies’ findings. (18,19), While RBCs count, Hematocrit, Hb, and female Platelets count values appeared to be higher in this study. MC V and WBCs were found to be lower than Western values. The comparison of our finding and other studies of hematological values in some african countries were presented in Table (8). The median values for RBCs count, hemoglobin, and red cell indices (HCT, MCV, MCH, and MCHC) as well as WBCs count greet similarity to the median values have been made by other African (10,11,12,13,14,15 and 16) While the platelet count values in this study appeared to be higher than in other African studies.

The reference intervals in this study varied slightly from the only previous made study in Asmara (Table 9). The lower normal limit of RBCs, MCHC, RDW, male (Hb, PCV, and PLATELET), and female WBCs RIs in this study was higher than in the study made by Nijat et al’s(5). The lower normal limit of females for (Hb, PLATELET, PCV, MCV, MCH) and male WBCs RIs in this study was lower than in Nijat’s study. The upper normal limit of male RBC, MCH, Hb, PCV, MCHC, and PLATELET RIs in this study was higher than Nijat study. The upper limit of females for Hb, MCH, PCV, MCV, and RDW RIs in this study was lower than in Nijat’s study.

In the present study we found that with an increase in age, there is a significant increase in BASOPHILS, RDW, and RDW-SD was observed. However, with an increase in age, a significant decrease in RBC, HB, HCT, and PLATELET was observed. Similar results have been reported by Isabel et al (21), MCV, MCH, and RDW showed a significant positive correlation with age in their study, whereas hemoglobin, hematocrit, RBC, MCHC, and platelet count showed a significant negative correlation. The amount of growth hormone secreted decreases with age. It is widely assumed that growth hormone stimulates erythropoiesis by increasing oxygen consumption of tissues and thus promoting tissue hypoxia, which in turn accelerates erythropoietin production by the kidneys. (22)

In our study, a positive significant correlation was observed between height and MONOCYTES, RBC, HB, HCT, MCH, and MCHC. On the other hand, a decrease in RDW-SD was observed with an increase in height. In previous studies, a significant negative correlation was reported between height and platelet count (23) and white blood cell count (23) On pubmed search we did not find any study which explored the relationship of RBCs, Hb, MCV, MCH, and MCHC with height

In our study, we found that MONOCYTES is negatively correlated with weight, depicting a decrease in MONOCYTES with an increase in weight. Whereas WBC, BASOPHILS, RBC, HB, and HCT were positively correlated with weight. Our finding contradicts Sephal et al (24)who found significant negative correlation between Hb and weight and height

In this study, a positive significant increase in BMI and BASOPHILS was observed with increase in BMI. However, negative correlation was observed between MONOCYTES and BMI. The remaining thirteen hematological parameters were not significantly correlated with BMI. Our results agreed with Mitraet al(25) study that reported a poor positive correlation between BMI and hematocrit WBCs and platetets among participants on other hand Farhangi et al (26) found that the levels of platelet counts were significantly higher among obese participants. Contrary to our findings Han et al (27) study show a positive correlation between WBC count and BMI

In our study, we found HB and HCT were significantly related to DBP. A significant positive relationship was observed between Hb, HCT, and DBP. Only three hematological parameters were found to have a significant correlation with SBP, namely, BASOPHILS, HB, and PLATELET. Out of the three parameters, a significant decrease in PLATELET was observed with an increase in SBP. However, a significant increase in BASOPHILS and Hb was observed with an increase in SBP. Jacob et al (28) found a significant increase in both systolic and diastolic blood pressure with higher values of hematocrit, hemoglobin, and red blood count. On the other hand, the findings of Kun Yang et al (29) show a negative relationship between PLATELET count with DBP.

In this study, we found that males had significantly higher levels of MONOCYTES, EOSIN, RBC, HB, HCT, MCV, MCH, and MCHC than females. The causes of these differences have been attributed to factors such as the androgen hormone’s influence on erythropoiesis and menstrual blood loss in females.(30) On the other hand PLATELET was higher in females than in males. The higher platelet count in females from puberty could be explained by estrogen promoting platelet production. (31)

## Conclusion

This study allowed us to establish the reference intervals of the complete blood count for healthy adults Eritrean in Asmara.

The hematological reference values for healthy adults in Asmara established in this study differ considerably from the reference values recommended by the Eritrean ministry of health

Gender, height, weight, BMI, and blood pressure parameters have all been shown to influence hematological parameters. More research is needed to establish hematological reference values for children, adolescents, the elderly, and pregnant women in order to avoid diagnostic errors and allow clinicians to more precisely interpret hematological examinations and improve the quality of medical care provided to patients.

## Data Availability

All data produced in the present study are available upon reasonable request to the authors

## Acknowledgements

The authors thank the Zone and subzone administrators and all study participants. Our thanks also go to the director of National Health laboratory, the head and staff of hematology department

## Reference

1. Ridley J. Essentials of clinical laboratory science. Nelson Education; 2010.

2. Boyd JC. Defining laboratory reference values and decision limits: populations, intervals, and interpretations. Asian J Androl. 2010;12(1):83. DOI: 10.1038/aja.2009.9

3. Iftikhar R, Khan NU, Iqbal Z, Kamran SM, Rodeheffer RJ. Haematological Parameters in Different African Populations: An Experience From United Nations Level 3 Hospital. Pakistan Armed Forces Med J. 2017;67(6):1068–72.

4. Wayne P. Defining, establishing and verifying reference intervals in the clinical laboratory; approved guideline. Clin Lab Stand Inst. 2008;

5. Siraj N, Issac J, Anwar M, Mehari Y, Russom S, Kahsay S. Establishment of hematological reference intervals for healthy adults in Asmara. BMC Res Notes [Internet]. 2018;1–6. Available from: https://doi.org/10.1186/s13104-018-3142-y DOI: 10.1186/s13104-018-3142-y

6. Https://worldpopulationreview.com/countries/eritrea-population.No Title.

7. Harris EK, Rodeheffer RJ. On dividing reference data into subgroups to produce separate reference ranges. Clin Chem. 1990;36(2):265–70. https://doi.org/10.1093/clinchem/37.9.1580

8. Harris EK, Wong ET, Shaw Jr ST. Statistical criteria for separate reference intervals: race and gender groups in creatine kinase. Clin Chem. 1991;37(9):1580–2.

9. Landis JR, Rodeheffer RJ. The measurement of observer agreement for categorical data. Biometrics. 1977;159–74. https://doi.org/10.2307/2529310

10. Osei-Bimpong A, McLean R, Bhonda E, Rodeheffer RJ. The use of the white cell count and haemoglobin in combination as an effective screen to predict the normality of the full blood count. Int J Lab Hematol. 2012;34(1):91–7. DOI: 10.1111/j.1751-553X.2011.01365.x

11. Awad K, Bashir A, Osman AA, Rodeheffer RJ. Reference Values for Hemoglobin and Red Blood Cells Indices in Sudanese in International Journal of Health Sciences and Research Reference Values for Hemoglobin and Red Blood Cells Indices in Sudanese in Khartoum State. 2019;(January).

12. Taha EH, Elshiekh M, Alborai A, Hajo EY, Hussein A, Awad KM, et al. Normal range of white blood cells and differential count of Sudanese in Khartoum state. 2018;5(4):784–7. https://dx.doi.org/10.18203/2349-3933.ijam20183116

13. Enawgaw B, Birhan W, Abebe M, Terefe B, Rodeheffer RJ. Haematological and immunological reference intervals for adult population in the state of Amhara, Ethiopia. 2018;23(7):765–73. DOI: 10.1111/tmi.13071

15. Dosoo DK, Kayan K, Adu-gyasi D, Kwara E, Ocran J, Osei. K, et al. Haematological and Biochemical Reference Values for Healthy Adults in the Haematological and Biochemical Reference Values for Healthy Adults in the Middle Belt of Ghana. 2012;(May 2014). DOI: 10.5402/2011/736062

16. Kueviakoe IM, Segbena AY, Jouault H, Vovor A, Imbert M. Hematological reference values for healthy adults in Togo. ISRN Hematol. 2011;2011.

17. Samaneka WP, Mandozana G, Tinago W, Nhando N. Adult Hematology and Clinical Chemistry Laboratory Reference Ranges in a Zimbabwean Population. 2016;(December). DOI: 10.1371/journal.pone.0165821

18. Kibaya RS, Bautista CT, Sawe FK, Shaffer DN, Rodeheffer RJ. Reference Ranges for the Clinical Laboratory Derived from a Rural Population Reference Ranges for the Clinical Laboratory Derived from a Rural Population in Kericho, Kenya. 2008;(February). DOI: 10.1371/journal.pone.0003327

19. Miri-dashe T, Osawe S, Tokdung M, Daniel N, Choji RP, Mamman I, et al. Comprehensive Reference Ranges for Hematology and Clinical Chemistry Laboratory Parameters Derived from Normal Nigerian Adults. 2014;9(5). DOI: 10.1371/journal.pone.0093919

20. Mohamed A, Eldin M, Hussien M, Badi R, Rodeheffer RJ. Reference ranges of white blood cells and platelets counts among Sudanese young adult males in Khartoum state. 5544(table 1):9–11. DOI:10.18088/ejbmr.3.3.2017.pp9-11

21. Isabel B, Nicoleta NM, José R, Garzón D, María C-I, Bessy B, et al. Evaluation of biochemical and hematological parameters in adults with Down syndrome. Sci Reports (Nature Publ Group). 2020;10(1). doi.org/10.1038/s41598-020-70719-2

22. Bijlani RL. Understanding medical physiology: a textbook for medical students. Jaypee; 2003.

23. Shimizu Y, Sato S, Koyamatsu J, Yamanashi H, Nagayoshi M, Kadota K, et al. Possible mechanism underlying the association between height and vascular remodeling in elderly Japanese men. Oncotarget. 2018;9(8):7749. DOI: 10.18632/oncotarget.23660

24. Acharya S, Patnaik M, Mishra SP, Rodeheffer RJ. Correlation of hemoglobin versus body mass index and body fat in young adult female medical students. Natl J Physiol Pharm Pharmacol. 2018;8(10):1371–3. doi: 10.5455/njppp.2018.8.0619912062018

25. Zarrati M, Aboutaleb N, Cheshmazar E, Shoormasti RS, Razmpoosh E, Nasirinezhad F. The association of obesity and serum leptin levels with complete blood count and some serum biochemical parameters in Iranian overweight and obese individuals. Med J Islam Repub Iran. 2019;33:72. doi: 10.34171/mjiri.33.72

26. Farhangi MA, Keshavarz S-A, Eshraghian M, Ostadrahimi A, Saboor-Yaraghi A-A. White blood cell count in women: relation to inflammatory biomarkers, haematological profiles, visceral adiposity, and other cardiovascular risk factors. J Health Popul Nutr. 2013;31(1):58. doi: 10.3329/jhpn.v31i1.14749

27. Han SN, Jeon KJ, Kim MS, Kim H-K, Rodeheffer RJ. Obesity with a body mass index under 30 does not significantly impair the immune response in young adults. Nutr Res. 2011;31(5):362–9. DOI: 10.1016/j.nutres.2011.04.002

28. Plange-Rhule J, Kerry SM, Eastwood JB, Micah FB, Antwi S, Rodeheffer RJ. Blood pressure and haematological indices in twelve communities in Ashanti, Ghana. Int J Hypertens. 2018;2018. doi: 10.1155/2018/5952021

29. Yang K, Tao L, Mahara G, Yan Y, Cao K, Liu X, et al. An association of platelet indices with blood pressure in Beijing adults: applying quadratic inference function for a longitudinal study. Medicine (Baltimore). 2016;95(39). DOI: 10.1097/MD.0000000000004964

30. Kibaya RS, Bautista CT, Sawe FK, Shaffer DN, Sateren WB, Scott PT, et al. Reference ranges for the clinical laboratory derived from a rural population in Kericho, Kenya. PLoS One. 2008;3(10):e3327. DOI: 10.1371/journal.pone.0003327

31. Daly ME. Determinants of platelet count in humans. Haematologica. 2011;96(1):10. doi: 10.3324/haematol.2010.035287

